# Exploring Patient and Staff Experiences of Video Consultations During COVID-19 in an English Outpatient Care Setting: Secondary Data Analysis of Routinely Collected Feedback Data

**DOI:** 10.1101/2020.12.15.20248235

**Authors:** HL Bradwell, RL Baines, KJ Edwards, SG Stevens, K Atkinson, E Wilkinson, A Chatterjee, RB Jones

## Abstract

**Background:** Video consultations (VCs) were rapidly implemented in response to COVID-19, despite modest progress prior to the pandemic.

**Objectives:** To explore staff and patient experiences of VCs implemented during COVID-19, and use feedback insights to support quality improvement and service development.

**Methods:** Secondary data analysis was conducted on 955 (22.6%) patient responses and 521 (12.3%) staff responses routinely collected following a VC between June-July 2020 in a rural, aging and outpatient care setting at a single NHS Trust. Patient and staff feedback were summarised using descriptive statistics and inductive thematic analysis and presented to Trust stakeholders.

**Results:** Most (93.2%) patients reported having ‘good’ (n=210, 22.0%), or ‘very good’ (n=680, 71.2%) experience with VCs and felt listened to and understood (n=904, 94.7%). Most patients accessed their VC alone (n=806, 84.4%), except for those aged 71+ (n=23/58, 39.7%), with ease of joining VCs negatively associated with age (*P*<.001). Despite more difficulties joining, older people were most likely to be satisfied with the technology (n=46/58, 79.3%). Both patients and staff generally felt patients’ needs had been met (n=860, 90.1%, n=453, 86.9% respectively), although staff appeared to overestimate patient dissatisfaction with VC outcome (*P*=.021). Patients (n=848, 88.8%) and staff (n=419, 80.5%) generally felt able to communicate everything they wanted, although patients were significantly more positive than staff (*P<*.001). Patient satisfaction with communication was positively associated with technical performance satisfaction (*P*<.001). Most staff (89.8%) reported positive (n=185, 35.5%), or very positive (n=281, 54.3%) experiences of joining and managing a VC. Staff reported reductions in carbon footprint (n=380, 72.9%) and time (n=373, 71.6%). Most (n=880, 92.1%) patients would choose VCs again. Inductive thematic analysis of patient and staff responses identified three themes: i) barriers including technological difficulties, patient information and suitability concerns; ii) potential benefits including reduced stress, enhanced accessibility, cost and time savings; and iii) suggested improvements including trial calls, turning music off, photo uploads, expanding written character limit, supporting other internet browsers and shared interactive screen. This routine feedback, including evidence to suggest patients were more satisfied than clinicians had anticipated, was presented to relevant Trust stakeholders allowing improved processes and supporting development of a business case to inform the Trust decision on continuing VCs beyond COVID-19 restrictions.

**Conclusions:** Findings highlight the importance of regularly reviewing and responding to routine feedback following the implementation of a new digital service. Feedback helped the Trust improve the VC service, challenge clinician held assumptions about patient experience and inform future use of VCs. The feedback has focussed improvement efforts on patient information, technological improvements such as blurred backgrounds and interactive white boards, and responding to the needs of patients with dementia, communication or cognitive impairment or lack of appropriate technology. Findings have implications for other health providers.

## Introduction

COVID-19 is a global health concern [1] that has resulted in the rapid implementation and digitalisation of many healthcare services [2,3]. As a result, video consultations (VCs) also referred to as remote, or virtual consultations [4] now form an integral part of both primary and outpatient (ambulatory) care. While limiting viral exposure and reducing potential risk of infection for patients and staff [4,5], VCs may also enable additional visual cues beyond the capabilities of telephone consultations, helping to further facilitate therapeutic relationships and experiences of care [2].

Although there is guidance on how to deliver VCs [6,7,8] and growing evidence exploring the rapid implementation of VCs in various areas of health care [4,9-17], there are relatively few empirical studies of VCs. For example, Doraiswamy et al. reviewed 543 articles relating to telehealth (including telephone, VC and other communication methods) during COVID-19, only 12% of articles presented empirical work, with few studies conducted in the United Kingdom (UK) focussing on VC [16]. Similarly, other research has focused on a single service such as orthopaedics or mental health [4,13,14]. While Dhahri et al.’s [15] research focuses on feedback across a range of outpatient services from 45 patients and 79 clinicians, this was only over a three week period.

The emerging research on remote healthcare and VC implementation seems to show some benefits. For example, VCs are seen as useful for social distancing [12], may provide quicker consultation times [4], reduce travel time for patients [14] and allow for safeguarding and risk assessment [13]. However, the research to date also shows areas of concern, such as technology limitations (impairing video/sound) [4,9,15], additional burden, lack of physical examination [9], low technology confidence and limited set-up support [4], further to impaired therapeutic interactions and reduced depth of clinical encounters [14].

However, as suggested above, the sample size of VC respondents in studies to date is arguably limited. Furthermore, no studies to the researchers’ knowledge have been reported from a British rural and deprived region with an aging/older population [18]. Documented implications of routine feedback on practice have also been lacking in previous literature. Our study thus aimed to investigate National Health Service (NHS) staff and patient experiences of the Attend Anywhere VC in Cornwall using routine feedback from a large sample and explore the impact of insights shared when presented to key stakeholders in the service.

## Methods

### Design

Secondary analysis of routinely collected anonymised survey data following a VC that was designed and distributed by the partner health care provider (NHS Trust); subsequent follow-up to disseminate results and assess impact of feedback with the Trust.

### Setting

The Cornwall Foundation Partnership NHS Trust provides mental health and community services for Cornwall and the Isle of Scilly (CIoS), a geographically isolated peninsula in the South West of England that experiences higher than average levels of deprivation [19]. By UK definitions, CIoS is very rural with 40% of the population living in remote areas [19], with an older age profile [24], and a single acute hospital located in the centre of the county. The Trust’s services include children’s and adolescent mental health, adult mental health and physical health, including but not limited to: learning disabilities, cardiac services, bladder and bowel, complex care and dementia, eating disorders, personality disorders, psychiatric liaison, palliative care, stroke nursing, speech and language therapy, diabetes, epilepsy, minor injuries, musculoskeletal, neuro-rehabilitation, physiotherapy, podiatry and respiratory nursing. This study was carried out by an independent research team using the anonymous data provided by the Trust.

### Video consultations

The Trust started using Attend Anywhere, a telemedicine platform for outpatient care [20,21], from 6^th^ April 2020. The implementation process followed guidance provided by NHS England and NHS Improvement [22]. Some appointments were still carried out face-to-face but many patients were offered VC or telephone contact during the study period.

### Data Collection

The Trust set up a system of routine feedback using an online survey (Appendix 1) based on the standardised feedback survey questions provided in the national guidance [22]. All staff and patients were invited immediately after participating in a VC to complete the online survey. To our knowledge, the survey was not designed with patient or public involvement due to the rapid implementation process. The rapid rollout of this feedback process may also be responsible for some limitations of the surveys themselves, including limitations in response options and understanding of the sample (for example, number of unique individuals and service accessed by patient). However, we have confidence that the data items used in the analysis are robust based on their recommended use by NHS England and NHS Improvement.

### Participants

Participation was voluntary, patient participants were patients or their carers. Survey respondents gave consent for their data to be used but some chose to not have comments publicly shared. Their data were included in analysis and production of the themes but their quotes are not included. We used data collected during the early implementation of VCs in response to COVID-19 (June 1^st^ – July 31^st^ 2020). During this time, 4234 Attend Anywhere appointments were completed. The feedback data used was from 955 patient and 521 staff responses out of the 4234 completed video-consultations. Sample size was largely pragmatic using data from as many patients (nearly 1000) that was thought possible to thematically analyse in a timely manner to give feedback to staff, during a two month period after an initial ‘settling down’ of the system but early enough to have practical use in assessing the utility of the method.

### Data Analysis

Descriptive statistics are reported for numerical data. Free-text responses were analysed by four researchers (HB, RB, KE, SS) using inductive thematic analysis [23]. Staff and patient comments were analysed separately initially. A comprehensive coding framework was developed. Patient and staff codes demonstrated high comparability and thus are presented as combined themes across the dataset, with areas of discordance discussed. Thematic analysis was selected as a useful and flexible method to generate a rich, yet detailed and complex account of qualitative data [23]. Adopting an inductive approach also helped to ensure identified themes arose from the data generated, as opposed to predefined concepts or ideas.

### Documented impact of routine feedback on real-world practice

Following analysis of results, summary presentations were created and presented to relevant stakeholders as rapid feedback between November 24^th^ 2020 and February 21^st^ 2021. Presentations were given in partnership with a service-user consultant to patients, patient representatives and professionals at an Experiences of Care Collaborative (ECCo) meeting within thecare system, the South West Outpatient Transformation group, the region’s Video Consultation forum, local online research dissemination events and to National audiences through the Outpatient Transformation regional leads meeting. Results were also shared with interested international healthcare providers (Finland).

### Patient and public involvement

The research question and study has been informed by patient input (through inclusion of patient experience) and a service-user consultant contributed towards reviewing and co-authoring the manuscript.

### Ethics

Ethical approval to conduct this secondary analysis was given by the Health Research Authority (HRA) and Health and Care Research Wales (HCRW), IRAS ID 286543 (27.07.2020). This manuscript has been prepared using SQUIRE reporting guidelines [24].

## Results

The results are presented in three sections: (i) quantitative results including participant characteristics; (ii) qualitative results; (iii) documented impact of this routine feedback on real-world practice. The Appendix gives further details on questions presented to staff and patients in the survey.

### Quantitative results

#### Patient age and device use

Just under a quarter (n=955, 22.6%) of the 4234 patient VCs resulted in feedback responses. As the data are anonymous, it was not possible to know if some individuals had completed the survey more than once in this two month period. Each data entry is therefore treated as an individual episode. The highest number of survey responses was received from individuals aged 31-50 years, with the lowest response from patients aged over 71. Half the patients used a laptop to access their VC. Device use varied by age (χ^2^= 68.9; 12df; *P*<.001); patients over 50 were much less likely to use mobile and more likely to use a tablet (Table 1).

**Table 1.** Use of devices for the video consultation by age group, showing numbers (percentages).

| AGE GROUP | DEVICE |  |  |  | TOTAL |
| --- | --- | --- | --- | --- | --- |
|  | Laptop | Mobile | Tablet | Other |  |
| <18 | 82 (50.3) | 44 (27.0) | 32 (19.6) | 5 (3.1) | 163 (17.1) |
| 18-30 | 76 (52.1) | 35 (24.0) | 24 (16.7) | 11 (7.5) | 146 (15.3) |
| 31-50 | 167 (50.2) | 87 (26.1) | 64 (19.2) | 15 (4.5) | 333 (34.8) |
| 51-71 | 120 (49.6) | 20 (8.3) | 70 (28.9) | 32 (13.2) | 242 (25.3) |
| >71 | 36 (62.1) | 1 (1.7) | 16 (27.6) | 5 (8.6) | 58 (6.1) |
| Total | 487 (50.9) | 189 (19.8) | 209 (21.9) | 70 (7.3) | 942 # |

#### Staff Characteristics

In total, 521 staff responses were received (Table 2), a response rate of 12.3%. The largest number of responses by profession were from Allied Health Professionals. The largest number of responses by department was from Community Mental Health. Most staff responses were completed following VCs with a patient for whom staff members had 1-3 prior contacts. Staff data, like patient data, are episodes rather than individuals.

**Table 2:** Staff respondents showing profession and department (n=521)

| Profession | N | % | Department | N | % |
| --- | --- | --- | --- | --- | --- |
| Nurse | 81 | 15.5 | ACS<sup>1</sup> Inpatient | 17 | 3.3 |
| Psychologist | 117 | 22.5 | ACS<sup>1</sup> Community | 114 | 21.9 |
| AHP<sup>4</sup> | 155 | 29.9 | Mental Health Community | 188 | 36.1 |
| Doctor | 61 | 11.8 | Mental Health Inpatient | 4 | .8 |
| Other | 107 | 20.5 | CAMHS<sup>2</sup> | 127 | 24.4 |
|  |  |  | Children's Services | 56 | 10.7 |
|  |  |  | Complex Care and Dementia | 10 | 1.9 |
|  |  |  | AMH<sup>3</sup> and Learning Disabilities | 5 | 1 |
| Total | 521 |  |  | 521 |  |

1. ACS: Adult community services (e.g. podiatry, spinal, physical, rehabilitation) 2. CAMHS: Child and adolescent mental health services 3. AMH: Adult mental health and learning disabilities 4. AHP: Allied Health Professionals

#### Patient overall experience and future intention

Most (n=890, 93.2%) patients reported having a ‘good’ (n=210, 22.0%), or ‘very good’ (n=680, 71.2%) overall experience of VC. A small number of patients had a ‘poor’ (n=13, 1.4%), or ‘very poor’ experience (n=17, 1.8%). Future intention could also be seen as a measure of satisfaction with VC; nine out of ten patients were ‘very likely’ (n=704, 73.7%) or ‘somewhat likely’ (n=176, 18.4%) to choose a VC in the future. Very few patients (n=28, 2.9%) suggested they were ‘somewhat unlikely’ (n=17, 1.8%) or ‘very unlikely’ (n=11, 1.2%) to use VCs in the future. Within the results, we were able to look at two aspects of overall satisfaction: satisfaction with the technology (video and sound) and satisfaction with the communication (more related to the clinician’s performance in this situation).

#### Patient and staff technical satisfaction

Three-quarters of patients reported having a ‘very positive’ experience of sound and video quality (n=732, 76.6%, n=728, 76.2% respectively). When combined, 67.6% (n=646) had a very positive experience of both video and sound (Table 3). Three-quarters of staff also reported a ‘positive’ or ‘very positive’ experience of sound and video quality (n=411, 78.9%, 76.5% respectively).

**Table 3.**
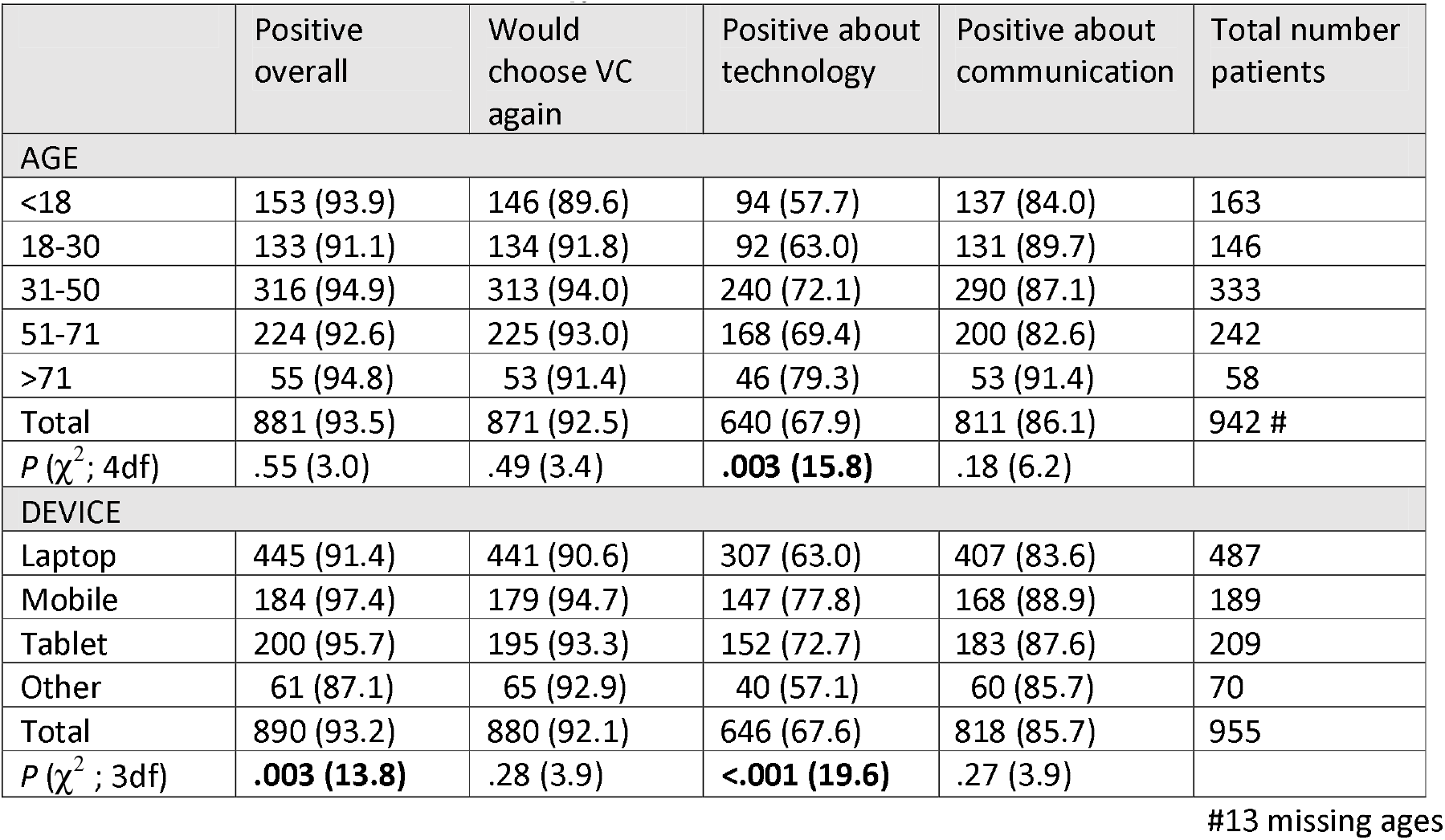
Four indicators of patient satisfaction with video-consultation shown by patient age and by device used, showing p-value from χ^2^test. P-values less than .05 in bold.

|  | Positive overall | Would choose VC again | Positive about technology | Positive about communication | Total number patients |
| --- | --- | --- | --- | --- | --- |
| **AGE** |  |  |  |  |  |
| <18 | 153 (93.9) | 146 (89.6) | 94 (57.7) | 137 (84.0) | 163 |
| 18-30 | 133 (91.1) | 134 (91.8) | 92 (63.0) | 131 (89.7) | 146 |
| 31-50 | 316 (94.9) | 313 (94.0) | 240 (72.1) | 290 (87.1) | 333 |
| 51-71 | 224 (92.6) | 225 (93.0) | 168 (69.4) | 200 (82.6) | 242 |
| >71 | 55 (94.8) | 53 (91.4) | 46 (79.3) | 53 (91.4) | 58 |
| Total | 881 (93.5) | 871 (92.5) | 640 (67.9) | 811 (86.1) | 942 # |
| $P(\chi^2; 4df)$ | .55 (3.0) | .49 (3.4) | **.003 (15.8)** | .18 (6.2) | |
| **DEVICE** |  |  |  |  |  |
| Laptop | 445 (91.4) | 441 (90.6) | 307 (63.0) | 407 (83.6) | 487 |
| Mobile | 184 (97.4) | 179 (94.7) | 147 (77.8) | 168 (88.9) | 189 |
| Tablet | 200 (95.7) | 195 (93.3) | 152 (72.7) | 183 (87.6) | 209 |
| Other | 61 (87.1) | 65 (92.9) | 40 (57.1) | 60 (85.7) | 70 |
| Total | 890 (93.2) | 880 (92.1) | 646 (67.6) | 818 (85.7) | 955 |
| $P(\chi^2; 3df)$ | **.003 (13.8)** | .28 (3.9) | **<.001 (19.6)** | .27 (3.9) | |

#13 missing ages

#### Patient and staff satisfaction with communication

The vast majority of patients felt they had been listened to and understood (n=904, 94.7%); had had their needs met (n=860, 90.1%) and felt they had been able to communicate everything they wanted (n=848, 88.8%). Overall 85.7% (n=818) of patients rated all three aspects positively (Table 3).

Most staff (n=419, 80.5%) felt able to communicate everything they wanted to, although satisfaction was slightly lower than for patients (80.5% Vs 88.8%; χ^2^=19.4; df=1; *P*<.001). Staff perceptions of patients feeling their needs were met were generally positive, with 57% (n=297), and 29.9% (n=155), responding ‘yes’ and ‘yes partially’ respectively. Only 2% (n=19) of patients said their needs were not met, while 11.1% (n=58) of staff believed patients felt their needs were not met suggesting an apparent discrepancy.

#### Association between patient satisfaction with technology and communication

Patient satisfaction with communication was very strongly positively associated with satisfaction with technical performance (χ^2^=104.0; df=1; *P*<.001) (Table 4). Only 41/955 patients were less satisfied with the communication despite being satisfied with the technology. Conversely, 213/955 patients remained positive about the communication despite being less positive about the technology.

**Table 4.**
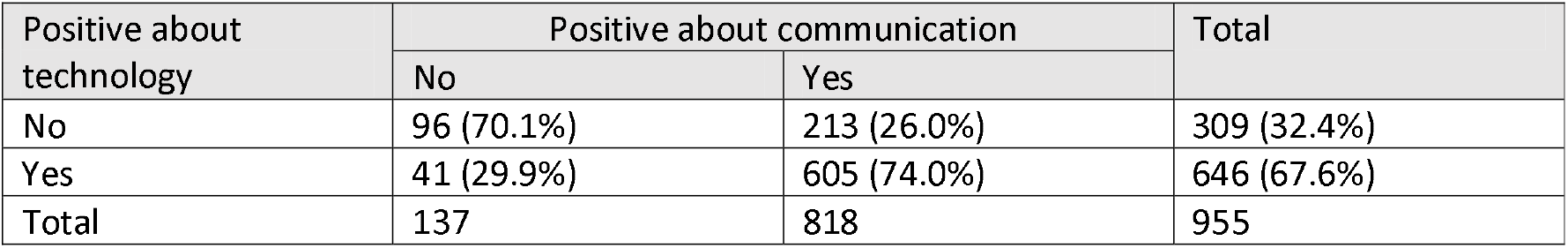
Crosstabulation of patients being positive about technology with being positive about communication.

#### Patient independence and accessibility

Most (n=806, 84.4%) patients stated that they could access their VC alone and that it was ‘very easy’ or ‘easy’ to join the VC call (n=849, 88.9%). However, more respondents over the age of 71 reported needing help (n=35/58, 60.3%) and fewer reported it easy to join compared to those under 31 (69.0% (n=40) Vs 92.6% (n=286) Vs; χ^2^=28.6, 3df, *P*<.001).

#### Influence of patient age and device used on outcome

The relationship between device use, age and satisfaction with technology and the VC was complex. On the one hand older patients were more positive about the technical experience, despite being more likely to need help accessing the VC and being less likely to find accessing the VC easy. Mobile users were also more positive (Table 3), and older people were less likely to be mobile users (Table 1).

#### Staff experience of managing and joining the call

Most staff respondents reported a ‘positive’ or ‘very positive’ experience when managing and joining the VC (n=466, 89.8%). A smaller number of staff responses indicated a ‘negative’ or ‘very negative’ experience (n=49, 9.4%).

#### Patient and staff perceived savings

Two thirds of patients reported a perceived saving in time (n=662, 69.3%), with more than half of respondents also reporting a perceived saving in money (n=544, 59.9%). There was no difference by age. Staff respondents most commonly identified a carbon saving (n=380, 72.9%) followed by time (n=373, 71.6%). Over a third of staff reported saving money (n=187, 35.9%). Just below a quarter of staff respondents reported a perceived saving on missed appointments or ‘did not attends’ (DNAs) (n=128, 24.6%). 24% (n=125) reported ‘other’ unspecified savings. Savings are explored in greater depth in the qualitative analysis below.

#### Patient versus staff perception

Overall, there appears to be good concordance between staff and patient feedback, with similar benefits noted for time and money savings. Mann-Whitney U tests demonstrated no significant difference between staff and patient ratings of video (n=1476, *P*=.152), or sound quality (n=1476, *P*=.768). However, significant differences between staff and patient responses were identified when reviewing whether patients had been able to communicate everything needed and feeling patient needs were met. On both occasions, staff responded more negatively than patients (n=1476, *P*<.001n=1476, *P*=.021 respectively). This could suggest staff overestimated patient dissatisfaction with VC outcomes or were not aware of patient experiences.

### Qualitative Results

#### Number of comments

One in eight (n=119/955) patients made 1384 free text comments in response to one or more of 16 questions (Appendix 1). Patients who rated their overall experience good or very good were much less likely to comment (11.8% Vs 43.1%, *P*<.001). Two thirds (n=350/521) of staff made 528 free text comments in response to one or more of the nine questions asked.

#### Overall themes

Inductive thematic analysis of free text responses identified three main themes: barriers, benefits and suggested improvements. While the overarching themes were the same based on staff and patient analysis, there was some variation in initial codes between the two groups, as discussed in the narrative below and demonstrated in Table 5. Unique identifiers are used for each quote, with S representing a staff comment and P representing a patient comment.

**Table 5:**
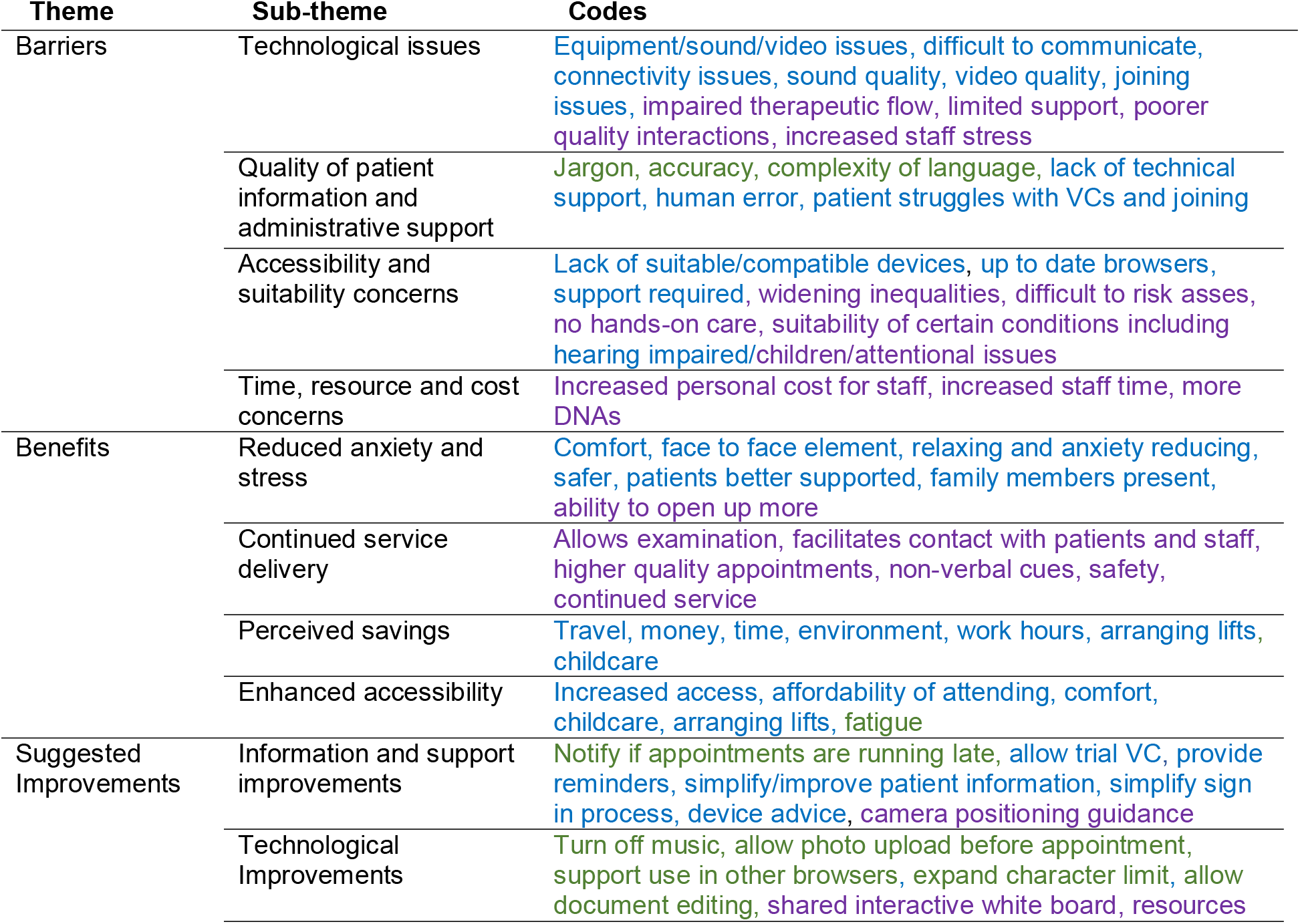
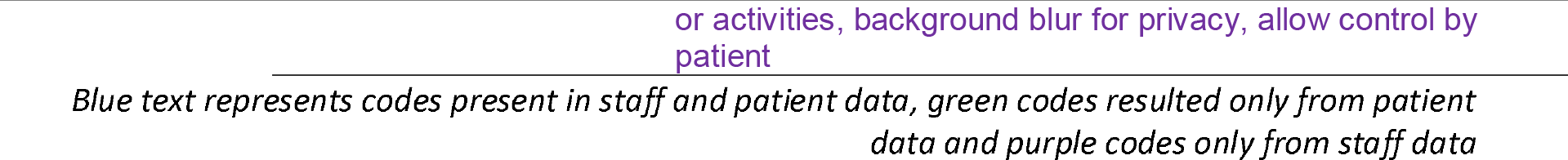
Qualitative patient feedback themes, sub-themes and codes.

#### Barriers

Participants related barriers from technological difficulties, quality of patient information and concerns about accessibility or suitability of using VC.

> Technology: Many patients identified concerns of connectivity, *“platform glitches”* (P94) or experienced delays between video and sound. For example, *“the picture kept freezing and pixelating”* (P741); *“the video quality is so poor it’s really hard to get much done”* (P332). In some cases, technical difficulties meant patients felt that their “*needs were not met”* (P305). Similarly, some clinicians reported having to *“resort to a telephone”* (S14) to supplement VC audio. Some staff reported VCs had *“a very negative effect on the quality of… therapy we can deliver”* (S28) and *“limit… the complexity of the conversation”* (S29) making it *“hard to pick up on body language”* (S144). For some staff, impairments caused by technological issues were seen as *“detrimental to patient care”* (S90), *“frustrating”* (S197, S409, S439) and *“stressful”* (S90). It is possible the connectivity issues seen in this study relate to the geographical character of the region of Cornwall, with staff reporting “*the general Cornish bandwidth is the obstruction here”* (S118). However, staff reported other platforms seemed to encounter less issues, suggesting the platform *“needs to be improved substantially and quickly”* (S22).
>
> Quality of Patient Information and Administrative Support: The quality of patient information, particularly joining instructions was repeatedly called into question by both patients and staff. One patient described the joining process as *“stupidly complex”* (P52). Others described being *“directed to a troubleshooting, jargon filled, suggestions page”* (P95). The accuracy of joining instructions was also questioned; *“the link doesn’t open as the instructions* [suggest]*”* (P236). Although a video version of the information was provided, this was described by participants as in need of further development and refinement to ensure inclusivity, particularly for “*deaf patients”* (P582). Similarly, staff felt the *“main issues”* (S75) for patients involved *“logging on”* (S71), *“the process of joining has lots of information to process”* (S75). The consequence being *“patients* [take] *a while to get into the appointment* [as] *the process* [is] *complicated*” (S150). Some clinicians needed to *“telephone and talk* [the patient] *through logging on”* (S286).
>
> Accessibility: Access to relevant devices, browsers, digital skills and/or confidence were also described as problematic by some participants. For example, *“unfortunately I was not up to date with technology”* (P319). Some patients reported needing to download alterative browsers or borrow other people’s devices due to “*outdated*” (P671) models. Staff also suggested *“it can be hard to engage those with limited IT equipment”* (S106). Similar to the quantitative findings outlined above, some participants reported needing help from family members or friends, as *“without* [them] *it would have been impossible”* (P540). Another *“96 year old had to pay for a carer to be present* [and] *2 hours of IT help from someone else”* (P47) raising further questions and concerns.
>
> Suitability concerns: While patient concerns focused mainly on technology and digital exclusion, staff had additional concerns of suitability based on patient illness or requirement. Some staff felt VCs were exacerbating *“health inequalities,”* for individuals with learning disabilities or living in residential homes as patients were *“often excluded from the review”* as computers were often located *“in an office”* (S202). This concern was echoed, as *“clients with learning disabilities”* often *“need reasonable adjustments to be facilitated to communicate”* (S15). The usefulness of VCs for dementia services was also queried, with the *“screen* [removing] *sensory aspects and visual clues*” (S130). VCs were also seen as unsuitable for dysphagia where *“a hands on approach is required to closely look, listen and feel as the person eats and drinks”* (S25). Patient conditions that may impede VC success as suggested by staff respondents also included *“cognitive, speech, language, fatigue, concentration, need for physical…assessments, environmental assessments, safety”* (S23). Other areas described as problematic included family therapy where the *“family had to sit side by side, so parents couldn’t see their child’s facial expressions”* (S278) and VCs with children generally, particularly with attentional needs, which *“meant* [the] *session was longer”* and less tasks were achieved than face-to-face (S136). Patient safeguarding and *“environmental assessments”* (S23) also appeared to be a key issue for clinicians; *“home visits remain hugely important to gather information to ensure patient safety”* providing more information *“such as how a person may be living and identify self-neglect, declines in functional skills or poor medication management”* (S130). Staff also suggested some conversations may *“be very challenging,”* over VC, such as discussing *“the risk of dying over the internet and not in person”* (S202) as some respondents felt *“discussing end of life care”* over video carries *“an increased risk of missing cues”* (S29). Patients raised concerns for people with hearing impairments as *“face to face is easier”* (P457) for lip readers due to video/sound delays, and character limitations in the VC chat function.
>
> Time, resource and cost ineffectiveness: A minority of staff reported concerns of time, resource and cost ineffectiveness, as while VCs “*saved a few minutes walking to the clinic and tidying the room for the assessment*” it could also “*add an extra appointment*” (S169) when a face-to-face consultation was needed. Some respondents also suggested VCs were longer due to a “*longer explanation time”* (S279) of how to long on etc. as described above. Technological issues also impacted duration, with patients *“having to change rooms*” (S198), “*change to telephone*” (S203) or “*re-join*” (S19) after disconnection. Furthermore, staff felt VCs “*take quite a bit of preparation prior to the consultation*” (S23), particularly in psychological services where staff may “*need to make electronic versions of therapy resources*” (S6). While 24.6% of staff reported reduced DNAs numerically, some staff suggested DNAs had increased; “*I’ve had more DNAs than when most of my visits were by car*” (S413). Interestingly, patients did not report time or resource ineffectiveness, and generally reported savings, as described below.

Interestingly, no patients described the therapeutic relationship or quality of care delivered by individual healthcare professionals as a barrier or limitation of VC use.

#### Benefits

Participants identified benefits for continued high quality service delivery, including reduced anxiety and stress, perceived savings, and enhanced accessibility. The majority of patients repeatedly described their VC experience as *“fantastic,” “excellent,” “amazing,” “wonderful,” “positive”* and *“useful”* (P558, P579, P153, P863, P600, P667). Participants also appeared to appreciate being able to have *“family members join the appointment”* (P15).

> Continued High Quality Service Delivery: For staff, VCs allowed them to “*continue to provide care*” (S47), “*maintain contact and show patients that we are here, that we are holding them in mind and we are motivated to help*” (S24). Clinicians noted without VCs, provision would be “*even more reduced*” (S47) and services such as “*psychotherapy*” have not needed to go “*on hold*” (S160). Staff who are “*shielding*” (S356) have also managed to maintain workloads, and also provide care for “*shielding patients”* (S394). Some clinicians reported good “*depth of therapy work*” which was “*less tiring than therapy by telephone*” (S60).
>
> Reduced Anxiety and Stress: Both patients and staff felt VCs were *“far less stressful”* (P796) than face-to-face. Several patients reported feeling *“more relaxed”* (P563) and less “*rushed”* (P392) as a result of avoiding certain stressors including arranging transport, arriving on time, finding and paying for parking and avoiding traffic. Reduced anxiety and stress was also reported among children, particularly for children who experienced anxiety about appointments or leaving the house. Clinicians noted child assessments could be supported by the presence of teaching staff further to parents alone, and for adults with learning disabilities, “*distant parents* [were] *able to join the review, whereas they wouldn’t have previously*” (S202). Staff also reported VCs were *“less stressful”* (S89), for the clinician themselves, “*increasing my wellbeing*” (S64) by “*saving stress*” (S209), “*anxiety and distress*” (S519). Other staff suggested that VCs “*can actually be more therapeutically productive*” (S170) with “*improved communication*” (S110) and “*better clinical contact*” (S200). Patients reported the removal of *“so much stress”* (P290) meant that they had *“more time to focus on what needs talking through”* (P290). Several participants suggested that because they *“didn’t feel so stressed”* (P695) and were in the comfort of their “*own home”* (P564), they were *“able to open up more”* (P695) often feeling more *“comfortable”* and *“relaxed”* (P564). Similarly, for staff, patients receiving care at home was seen as beneficial, such as for a “*post-natal mum*” who “*felt more comfortable in their own home*” (S126). VCs were also seen to reduce “*anxiety for patients concerned about face-to-face appointments due to COVID*” (S29) and allowed “*concordance*” (S58) in patient care.
>
> Perceived savings: Both patient and staff reported time, monetary and environmental savings. Some patients reported saving *“over £20 in transportation costs”* (P98) and being able to now ‘afford’ an appointment as a result of time and cost savings. For example, VCs *“have genuinely changed my life, …being accessible for my needs and being able to afford an appointment”* (P153). Travel and cost savings may be particularly prevalent in *“Cornwall”* where it *“is always difficult to travel for appointments”* (P262). Many staff also suggested VCs save “*time, money and travelling for all concerned*” (S324), also reducing “*carbon footprint*” (S75) by saving “*on travel*” (S315), “*paper*” (S242) and *“printing resources*” (S261). Many clinicians reported they could “*see patients more intensively*” (S7) and “*complete an increased number of appointments in a day*” (S19) with *“increased capacity as a whole”* (S17). Clients who were often “*late due to travel*” (S81) were now on time. Clinicians reported saving “*90 minutes in the car visiting a patient who is just as happy to be seen by video*” (S307). Some staff also reported that they “*rarely get DNAs*,” and that VCs “*must have saved* [The Trust] *a lot of money*” (S444).
>
> Enhanced accessibility: Although digital exclusion was thought to reduce accessibility for some as outlined above, the majority of free text responses suggested VCs facilitated service accessibility in a number ways. Firstly, due to certain conditions and reduced mobility, some respondents found “*trips to the hospital very tiring and difficult”* (P20). VCs removed this experience. Childcare and employment cost savings were also described helping to increase accessibility. For example, one participant suggested for a face-to-face appointment they *“would have been dragging all three kids along for what ended up being something that the video call was able to address”* (P863). The *“option of video call”* was also considered *“useful”* for those who are employed or *“working parents”* (P335). Patients could schedule weekly appointments around their employment, something that is not always possible when relying on face-to-face appointments. Furthermore, a number of patients who are *“not able to drive”* or *“can’t drive”* described VCs as *“much more convenient”* (P307, P846, P307) due to enhanced independence and removal of reliance on others. Similarly, staff reported increased accessibility for numerous patients who ordinarily “*cannot travel to appointments*” (S77) such as those with “*mobility issues*” (S201). The lesser time requirement for patients to attend via VC was considered to “*increased availability of services to clients with other commitments*” (S274), “*work engagements*” (S153) or “*caring responsibilities*” (S280).

As a result of the benefits encountered, many patients expressed a strong desire for VCs to be made available beyond the COVID-19 pandemic: *“I hope that all of our future appointments will be held this way”* (P863) highlighting an important element of patient choice.

#### Suggested improvements

Finally, staff and patients suggested improvements in two main areas information and technology.

> Information Improvements: Respondent suggestions included the simplification of patient guidance and information, device-specific advice and suggested device use for optimum VC experiences. For example, some patients reported positioning their *“iPad onto the floor, so I could show… my feet and me walking”* (P248). This would have been less feasible if using *“a desktop computer”* (P248). Other participants suggested it was *“a bit challenging getting* [the] *camera in position to demonstrate me doing the exercises”* (P809). Providing device specific information and recommending particular devices based on service requirements and availability may therefore be beneficial. Patients also requested some method of notification if clinics were running late.
>
> Technology Improvements: Related to platform functionality; patients requested the ability to *“turn off the music while sitting in the waiting room”* (P388) as it was considered *“terrible”* (P517) and repetitive by some. Patients also expressed a desire to be able to “*upload photos or videos”* (P20) to the consultation and an ability to *“change the mobile camera being used”* (P145). Further platform related improvements suggested included expanding its functionality to “*other browsers”* (P342) and having the character limit expanded beyond 200 characters for patients who need to use the text function. Other suggestions made by staff included more interactive screen sharing capabilities, as currently clinicians “*cannot see* [the] *client anymore*” (S6) when sharing their screen. Others thought it would be useful if the clinician could “*allow the client to take control of a particular programme you are sharing*” which could help with “*engaging children so you could play games together*” (S6). A similar desire was noted for adult Cognitive Behavioural Therapy, with staff requesting a “*white board*” (S119) to “*draw things out on such as CBT formulations*” (S52). The absence of such functionalities meant that diagrams were completed less “*collaboratively*” than if patients were “*in the room*” (S107). Finally, related to digital skills and confidence, some patients expressed a desire for a ‘practice run through’ to be made available so people could familiarise themselves with the technology prior to their consultation. Interestingly, no suggestions for healthcare training were proposed by participants although this may reflect the questions asked in the feedback survey. For enhanced security, staff requested a “*blurred background”* (S3) option “*to help protect privacy*” (S3) and “*improve confidentiality*” (S375). This seemed particularly relevant for clinicians who “*work with forensic clients*” (S3). An additional improvement would be having the ability to “*lock the room once everyone is in*” after experiencing “*incidents of other staff joining private, confidential therapy sessions uninvited*” (S8).

#### Comparison of Patient and Staff feedback

Generally, staff and patient responses showed high congruence, evidenced in the similarity of codes and subsequently combined themes. However, as with quantitative results, where more staff responded with concern about patients feeling their needs where met than patients themselves, the qualitative data also demonstrates some evidence of clinicians believing VCs impaired the therapeutic flow or produced poorer quality interactions.

Patients provided no indication of dissatisfaction with the clinician’s communication, outcomes or care received, other than issues resulting from the technology. While staff concerns on meeting patients’ needs also commonly resulted from technical issues, generic concerns were also shared on therapeutic quality and missing cues or body language via VC. Additionally, patients highly regarded the increased accessibility of healthcare, and found appointments less stressful. The codes on time, resource and cost ineffectiveness were provided only by staff, while patient data strongly supported perceived savings, across time, money, travel and environmental impact. Staff and patients showed similarity in the reported requirement for improved sound and video quality. Patients additionally requested the removal of waiting room music, expanding of character limits and trial runs, while staff requested blurred backgrounds, interactive shared screens. Both patients and staff requested simplified/improved patient information.

### Documented impact of routine feedback on real-world practice

Following the initial analysis of results, the authors presented the above findings to relevant patient and professional stakeholders to provide rapid feedback on barriers and facilitators, possible areas for improvement and ways to encourage the sustainable use of VCs. Findings were positively received by both clinicians and patients. In particular, clinicians reported underestimating patient satisfaction with VCs, and were surprised to see such high levels of satisfaction with the service and particularly patient perceptions of communication quality and feeling their needs were met. The presentations aided in revising clinician perceptions and some initial changes to VC practice have already been instigated, including replacement of waiting room music with bird song and efforts to improve patient information. The use of routine feedback and its analysis was thus instrumental in instigating some initial improvements for the use of VCs within the NHS Trust and promoting further conversations around future VC use and improvements. The implementing NHS Trust has now confirmed procurement of Attend Anywhere for VC service provision to continue beyond COVID-19 restrictions.

## Discussion

This research contributes towards existing literature exploring staff and patient experiences of the rapid rollout of VCs for outpatient care during the COVID-19 pandemic [20], with clear implications for policy, practice and future research. Specifically, this work contributes towards the VC evidence base with a considerably larger sample than prior evaluations, in a rural setting and with a focus on VCs rather than telephone. We also provide documentation of impact of routine feedback.

### Principal Results

The vast majority of patients rated their VC experience as ‘good’ or ‘very good,’ feeling listened to and understood, able to communicate everything they wanted to and feeling their needs had been met. Many patients reported saving time and money and over 90% would likely choose a VC in the future, although it remains unclear whether this resulted from no perceived ‘alternative’ option due to the global pandemic, or positive patient experiences and related motivations. Further exploration would be beneficial including analysing patient experience over time. Staff also generally supported use of VCs through positive experiences of joining and managing calls, being able to communicate all that was needed and feeling the patients’ needs had been met, although agreement on this latter issue was not unanimous. Staff also reported savings, mostly for carbon footprint and time. Qualitatively, both staff and patients noted increases in service accessibility and affordability. As a result, this Trust commissioned Attend Anywhere for future (COVID and post-COVID) use and other Trusts should consider making VCs a permanent option, beyond pandemic restrictions.

Patients aged 71+ were the only age group where a larger percentage of respondents reported needing support accessing their VC than those able to access alone. Nevertheless, there was no clear age gradient in satisfaction with VC. Indeed, older people were more likely to be positive about the technology than younger people. This may reflect higher expectations among younger people. While high levels of positive experience were reported across all device types, more users of tablet or mobile devices were positive. The relationship between age, device use, and positive experience of VC is complex. Research suggests the small size of mobile phones can pose a barrier for older adults, coupled with declining dexterity and vision, mobiles are mainly used by older adults for calls and texting [26], while tablet computers are more popular for online access among older people than younger people [27]. Older working age groups may be more likely to be prefer technologies they are familiar with at work (often desktops with poor cameras and sound systems), while older people new to computing use tablets as their ‘entry’ device [28].

Clinical areas of less suitability for VCs were also noted, particularly by staff. Further research is needed to identify when VCs work best, for who and in what context. While Greenhalgh and colleagues [7] have already provided guidance on appropriate and inappropriate use of VCs, a more granular understanding from both a patient and professional perspective may be required. Some limitations were noted for spinal services, neurology, for children, attentional issues and assessing dysphagia. The previous guidance [7] suggested inappropriate contexts included patients at high-risk, patients requiring internal examination and patients with challenges affecting ability to use technology. This suggestion is supported in our work, with staff suggesting VC was less appropriate for patients with learning disabilities, communication disorders, fatigue, cognitive issues or dementia.

Barriers described by staff and patients included technological difficulties, quality of patient information, administrative errors and accessibility or suitability concerns. Conversely, identified benefits included reduced stress and anxiety for patients and staff, the opportunity to ‘open up more’ for patients as a result of enhanced comfort, cost and time savings, increased sense of affordability and service accessibility. Finally, participants suggested a number of improvements such as simplifying patient information, notifications for late appointments, ability to turn off waiting room music, shared interactive white board, blurred backgrounds and ‘practice run’ opportunities to increase familiarity and digital skill confidence. Other suggested improvements include allowing photo or video upload to the appointment, swapping between cameras used, extending Attend Anywhere to other internet browsers and expanding the character restrictions of the chat function, particularly important for accessibility of deaf patients. This study thus has a number of practical implications. Routinely collecting and responding to feedback is likely to be an integral aspect of service improvement, as demonstrated through this work. Feasible improvements such as those reported here are likely to have important impacts on staff and patient experience.

The need to simplify and improve patient information was highlighted as a key barrier by both staff and patients and this may best be achieved in co-design with patients. While the implementation of VCs was a rapid response, actively involving patients and the public creating digital related information may improve accessibility, relevance and understanding. An implication of this work is thus an identified need to establish best practice for rapid co-design, when implementation timing is critical. Any future patient information may also include guidance for patients on camera positioning to reduce another barrier identified in this work. Healthcare services may also benefit from recommending particular devices based on their functionality and service requirements. For example, larger, static screens may be suitable for child therapies or family-based interventions where patients and families need to sit side-by-side. Alternatively, VCs including assessment of movement may be better suited to more portable devices such as mobiles or tablets.

An important consideration for VCs is safeguarding. Bhardwaj et al. [13], reported clinicians were confident in performing safeguarding and risk assessments remotely. Our results indicate otherwise, as clinicians reported home visits were key for patient safeguarding, to allow monitoring of self-neglect, decline in wellbeing or poor medication. Home visits for patients requiring environmental assessment could therefore be prioritised for face-to-face appointments, as could consultations relating to the identified, less appropriate contexts. However, this assumes environmental assessments are not possible via VC, where perhaps a visual tour of the home environment during a VC would suffice. Thus an alternative implication is guidance for clinicians to this regard. A further concern was raised by clinicians consulting with patients in residential care, with respect to widening health inequalities, as patients are often unable to attend consultations where computers were housed in staff offices. Additionally, some patients in these contexts appeared unable to appropriately position their camera, suggesting potential for solutions such as affordable telepresence devices or robotics in residential care to facilitate VCs. This could respond to two barriers i) that of patients being excluded from VCs due to equipment in staff offices and ii) challenges for appropriate camera positioning.

As the final practical implication, in response to patients noting administrative and human errors, the collaborative development of checklists and supportive training may be beneficial. Trusts could perhaps include on-screen checklists on patient records to ensure scheduling VCs is followed by provision of an appropriate link, patient facing guidance and set-up support. Clinicians may also consider promoting benefits of teachers or distant relatives attending a VC.

Our study also raises implications for the collection and use of routine feedback. The clinicians in our sample overestimated patient dissatisfaction with VCs. More clinicians also responded negatively to communication quality than patients. A minority of clinicians reported some impairment to therapeutic flow. Our presentations of these results to Trust stakeholders supported this observation, with clinicians surprised about high patient satisfaction. Some clinicians reported avoiding VCs for fear of patient dissatisfaction. The provision of this routine feedback thus aided in addressing staff perceptions. It is possible low staff expectations for VCs somewhat explains low documented uptake of VCs in comparison to telephone calls in previous research [10,13,14]. When collecting routine feedback, critically considering the purpose is important. For example, Sibley and colleagues recently likened the increasing collection of patient feedback as an “*avalanche… with experience now tracked, monitored and measured to an almost obsessive degree*” [31, p.4329]. However, to what end and for what purpose? Reflecting previously acknowledged concerns around the ethics of collecting patient feedback that leads to minimal direct benefit [32,33], Sheard et al. [34], suggested all patient feedback tools must be meaningfully usable by those providing frontline care, otherwise it becomes “*unethical to ask patients to provide feedback which will never be taken into account*” [34, p.51]. Service provides should thus ensure routinely collected feedback (including after an Attend Anywhere appointment) is meaningful for both patients and clinicians, serves a beneficial function beyond mandatory feedback collection and focuses on care delivery aspects most important to patients and clinicians. Future research may consider what feedback methods are most effective in encouraging responses, particularly in the new ‘digital norm,’ with staff members supported and empowered in acting upon and responding to feedback received.

### Limitations

The first limitation of this research is reliance on a self-selected sample of individuals who have attended or facilitated a VC and chosen to provide feedback. Experiences or barriers for those unable, or choosing not to use VCs currently remain unknown, as do experiences of those not providing feedback following their VC. Nevertheless, from the present study we know that many hundreds of staff and patients had a positive experience of VCs. Secondly, due to the anonymous nature of the data set, we were unable to identify how many individuals completed the survey on repeat occasions, thus results may be skewed by repeat respondents. Thirdly, this research relies on secondary data and the subsequent questions/scales used by the Trust. While informed by the “One week implementation guide for NHS Trusts and NHS Foundation Trusts” provided by NHS England and NHS Improvement [22], the survey has some limitations. Furthermore, to the researchers’ knowledge, the survey was not created in co-design with all relevant stakeholders including patients and carers. The questions asked may not therefore reflect the most important aspects of VC experience for patients. However, strengths of this research include the exploration of a relatively large data set of staff and patient experiences from an outpatient setting during the early stages of VC implementation. Unlike existing literature mainly focusing on primary care or specific health services, this research explored the use of VCs across outpatient services more broadly, helping to address limitations within existing COVID-response literature [4, 10].

Based on the limitations of this work and reliance on feedback from patients succeeding in accessing VCs, future research could explore challenges and barriers for excluded patients, particularly those considered ‘seldom heard’ or marginalised in the context of digital health [35]. Murphy et al. [12], noted previously that digital consultations increase access for those with IT skills, but may enforce existing health inequalities. Thus future research should explore accessibility, including; what works for who, when, and in what circumstances. This could also aid in supporting suitability guidance and ensure the full potential of VCs is realised. There is an important balance required between acknowledging increased accessibility for some patient groups through digitalisation (who may encounter difficulties accessing face-to-face services), while acknowledging a reduction in accessibility for others. This aspect needs urgent further work, and reiterates the importance of patient choice and availability of multiple mediums to access health and care services.

The nature of this study meant we were also unable to conclude on a number of additional factors, highlighting scope for further research. Apparent efficiency savings in service delivery via VC should be explored to assess the impact of VCs on clinician and patient experience. While intensive working may aid meeting growing healthcare requirements, a danger posed is workforce burnout. Related to efficiency, further research should look to establish DNAs prior to and since implementation of VCs, with almost a quarter of staff reporting less DNAs using VCs than usual practice. It would be interesting to explore if DNAs are linked to patient inability to access VCs or lack of confidence with technology. Other implications for future research include a need to identify the mechanisms responsible for the positive patient experiences and high levels of future VC use intentions as demonstrated by the Trust in this research. By doing so, other Trusts and healthcare services can engage with acknowledged areas of best practice. Economic evaluations that incorporate clinician, patient and environmental savings may also be beneficial, although it is important to emphasise that potential cost savings should not take precedence over patient safety, quality of care and stakeholder experience.

Additionally, suggestions made in this research that VCs improve ‘affordability’ of appointments and comfort in sharing personal or clinical information are important areas of future interest. Future research questions could include: how and in what ways, have video consultations affected patient accessibility? Similarly, how if at all, do video consultations affect the therapeutic relationship? Future research could compare patient satisfaction with more conventional face-to-face consultations or other VC platforms. Additionally, the range and type of consultations available to patients are currently limited and expressed satisfaction may reflect a lack of choice or alternatives. Future research may review patient experiences over time, particularly during times of heightened and reduced COVID-19 restrictions.

### Comparison with Prior Work

Generally, our results continue to demonstrate positive experiences for staff and patients of VC’s during the pandemic [4,9,10,16], furthering previous work with smaller samples and narrower focus. For example, congruent with Gilbert et al. [4], we found positively perceived aspects of VCs included reduced travel times and reduced impact of travel on symptoms such as fatigue [4]. Other similarities with prior work include low confidence reported among some participants [4], and negative impact of technological limitations and difficulties on patient experience [4,9,12,13]. Findings from this research regarding age differences in independent use and family involvement are also congruent with other research [12,29]. However, given the difficulties that many older people have in travelling to outpatient clinics [30] and the largely high acceptability of VC use reported in this current study for older people, no quick assumptions should be made about the unsuitability of VC for older people.

Areas of divergence with existing literature include patients reporting higher levels of satisfaction and willingness to use VCs in the future than in previous work [4]. Previous feedback was collected within an entirely orthopaedic service which could suggest greater satisfaction and use intentions are seen here due to the variety of services included, which may better translate to VC than orthopaedics, as perhaps a more ‘hands-on’ service. However, this would need further exploration, as survey limitations impair our understanding of exactly which service patient respondents accessed. Other contributions of this research include the identification of additional benefits including enhanced comfort and subsequent ability to ‘open up more.’ This contrasts with the results of Liberati et al. [14] who reported impairments to depth of conversation and relational quality via remote means. While this article reports on a larger sample, Liberati et al. [14] also reported on qualitative interviewing. This incongruence in results is therefore worth exploring further, perhaps across specific psychological therapy services. In contrast to Isautier et al. [9] suggesting telehealth limitations included poorer quality of communication, our results suggested the large majority of patients were satisfied with the VC aspects related to communication, with combined technical satisfaction being lower, congruent with Kayser et al. [10]. Here, we additionally provide further insight into the influence of patient age and device used in predicting overall VC experience, with implications for targeted consultations in future. This research also provides interesting insight into both staff and patients reporting increased sense of accessibility, and patient perception of enhanced affordability. Our work therefore contributes towards furthering previous research [4,9-17] reporting on small sample sizes and generally single service focus, while we report on a comparatively large sample across outpatient services, in a rural and aging setting [18,24].

## Conclusion

In conclusion, the majority of NHS staff and patients reported positive experiences of video consultations for outpatient care in a rural, aging and deprived setting. Patients often felt listened to, able to communicate their needs and understood, and staff and patients noted resource savings and enhanced accessibility. However, some barriers identified technological difficulties, accessibility of patient information and accessibility or suitability concerns require further attention if the potential benefits of VC are to be realised and use is to be sustained. Implications of this research include the implementation of patient suggested improvements, including trial calls, turning music off, facilitating photo uploads, expanding written character limit and supporting VCs on other browsers. Future work may explore accessibility and experience of patients excluded from this work, through lack of VC access. This work additionally demonstrated real-world impact of routine feedback, and raises further discussion on future use of routine staff and patient experience data.

## Data Availability

Data sharing statement: Not all patients have consented for their comments to be published, thus the full dataset is not available publicly, but interested parties may enquire with the authors for further details.

## Authors contributions

All authors read and approved the manuscript.

HB analysed and interpreted results and led on producing the manuscript.

RB analysed and interpreted results and substantively contributed towards the manuscript.

KE analysed and interpreted results and substantively contributed towards the manuscript. SS reviewed and contributed towards the manuscript.

KA provided expertise in patient experience and engagement, reviewed and substantively contributed towards the manuscript.

EW facilitated access to patient experience data, provided contextual data, reviewed and substantively contributed towards the manuscript.

AC reviewed and substantively contributed towards the manuscript.

RJ analysed and interpreted results, reviewed and substantively contributed towards the manuscript.

## Data sharing statement

Not all patients have consented for their comments to be published, thus the full dataset is not available publicly, but interested parties may enquire with the authors for further details.

## Appendices

## Appendix 1. Survey Questions and Response Options

Each question beginning with ‘COMMENT’ allowed for a free-text response. Patient survey is displayed first followed by staff survey.

### Patients

1. Please tick this box if you do not wish your comments to be made public
2. Service Line
3. Department
4. Team
5. Overall, how was your experience of our service very poor, poor, neither good nor poor, good, very good, don’t know
6. COMMENTS: Overall, how was your experience of our service?…
7. How easy was it to join the video appointment? very difficult, difficult, neither easy or difficult, easy, very easy
8. COMMENTS: How easy was it to join the video appointment?…
9. Joining the call very negative, negative, unsure, very positive
10. COMMENTS: Joining the call…
11. Sound quality very negative, negative, unsure, very positive
12. COMMENTS: Sound quality…
13. Video quality very negative, negative, unsure, very positive
14. COMMENTS: Video quality…
15. Being able to communicate everything you wanted to very negative, negative, unsure, very positive
16. COMMENTS: Being able to communicate everything you wanted to…
17. Feeling listened to and understood? very negative, negative, unsure, very positive
18. COMMENTS: Feeling listened to and understood?…
19. Feeling your needs were met very negative, negative, unsure, very positive
20. COMMENTS: Feeling your needs were met…
21. How likely are you to choose a video appointment as an option in your future care? very unlikely, somewhat unlikely, neither likely nor unlikely, somewhat likely, very likely
22. COMMENTS: How likely are you to choose a video appointment as an option…
23. Were you able to access this video appointment by yourself? yes, no, someone helped me
24. COMMENTS: Were you able to access this video appointment by yourself?…
25. What type of device did you use for your video appointment? laptop, mobile, tablet, other
26. COMMENTS: What type of device did you use for your video appointment?…
27. Which internet browser did you use for your video appointment? chrome, safari, other, not sure
28. COMMENTS: Which internet browser did you use for your video appointment…
29. What is your age? under 18 yrs, 19-30, 31-50, 51-71, 71 yrs +
30. COMMENTS: What is your age?…
31. COMMENTS: What specialty or service was your Video Appointment for?…
32. Has using this service helped to save you either time, money or other benefits? eg, travel fares, fuel, parking, lost earnings, Time, money, other
33. COMMENTS: Has using this service helped to save you either time, money…
34. COMMENTS: We are constantly looking at ways at improving our service

### Staff

1. Service Line
2. Department
3. Team
4. Channel
5. Profession:
6. Is using the video appointment system saving you any of the following: time, money, DNAs, carbon footprint, other 2
7. COMMENTS: Is using the video appointment system saving you any of the following: time, money, DNAs, carbon footprint, other
8. Have you met this patient face to face previously?
9. COMMENTS: Have you met this patient face to face previously?…
10. How many contacts have you had with this patient:
11. COMMENTS: How many contacts have you had with this patient:…
12. Managing and joining the call
13. COMMENTS: Managing and joining the call…
14. Sound quality
15. COMMENTS: Sound quality…
16. Video quality
17. COMMENTS: Video quality…
18. Being able to communicate everything you wanted
19. COMMENTS: Being able to communicate everything you wanted…
20. Did you feel the patient was able to communicate everything and had all of their needs met?
21. COMMENTS: Did you feel the patient was able to communicate everything..
22. COMMENTS: If there were issues please say how the experience could be improved

## References

1. Aprahamian I, Cesari M. Geriatric Syndromes and SARS-COV-2: More than Just Being Old. J Frailty Aging. 2020;9(3):127–129. doi: 10.14283/jfa.2020.17

2. Greenhalgh T, Wherton J, Shaw S, Morrison C. Video consultations for covid-19. BMJ. 2020;368:m998. doi: 10.1136/bmj.m998

3. Ohannessian R, Duong TA, Odone A. Global Telemedicine Implementation and Integration Within Health Systems to Fight the COVID-19 Pandemic: A Call to Action. JMIR Public Health Surveill. 2020;6(2):e18810. doi: 10.2196/18810

4. Gilbert AW, Billany JCT, Adam R, Martin L, Tobin R, Bagdai S, et al. Rapid implementation of virtual clinics due to COVID-19: report and early evaluation of a quality improvement initiative. BMJ Open Quality 2020;9(2):e000985. doi: 10.1136/bmjoq-2020-000985

5. Hollander JE, Carr BG. Virtually Perfect? Telemedicine for Covid-19. New England Journal of Medicine. 2020;382(18):1679–816. doi: 10.1056/NEJMp2003539

6. Iyengar K, Jain V K, Vaishya R. Pitfalls in telemedicine consultations in the era of COVID 19 and how to avoid them. Diabetes & Metabolic Syndrome: Clinical Research & Reviews. 2020;14(5), 797–799. doi: 10.1016/j.dsx.2020.06.007

7. Greenhalgh T, Shaw S, Seuren L, Wherton J. NHS, Video consultation information for NHS Trusts and Foundation Trusts. IRIHS Research Group. 2020. Available from: https://www.england.nhs.uk/coronavirus/wp-content/uploads/sites/52/2020/08/C0638-nhs-vc-info-for-nhs-trusts.pdf (Accessed 4th December 2020).

8. RCN. Remote Consultations Guidance Under COVID-19 Restrictions. Royal College of Nursing. (2020) https://www.rcn.org.uk/professional-development/publications/rcn-remote-consultations-guidance-under-covid-19-restrictions-pub-009256 (Accessed 20th November 2020).

9. Isautier JM, Copp T, Ayre J, Cvejic E, Meyerowitz-Katz G, Batcup C, et al. People’s Experiences and Satisfaction With Telehealth During the COVID-19 Pandemic in Australia: Cross-Sectional Survey Study. J Med Internet Res 2020;22(12):e24531. doi: 10.2196/24531

10. Kayser MZ, Valtin C, Greer M, Karow B, Fuge J, Gottlieb J. Video Consultation During the COVID-19 Pandemic: A Single Center’s Experience with Lung Transplant Recipients. Telemedicine and e-Health. 2020. doi: 10.1089/tmj.2020.0170

11. Barsom EZ, Feenstra TM, Bemelman WA. et al. Coping with COVID-19: scaling up virtual care to standard practice. Nat Med. 2020;26, 632–634. doi: 10.1038/s41591-020-0845-0

12. Murphy M, Scott LJ, Salisbury C, Turner A, Scott A, Denholm R, Lewis R, Iyer G, Macleod J, Horwood J. Implementation of remote consulting in UK primary care following the COVID-19 pandemic: a mixed-methods longitudinal study. British Journal of General Practice 8 February 2021; BJGP.2020.0948. doi: 10.3399/BJGP.2020.0948

13. Bhardwaj A, Moore A, Cardinal RN, Bradley C, Cross L, Ford TJ. Survey of CAMHS clinicians about their experience of remote consultation: brief report. BJPsych Open 2021;7:e34. doi: 10.1192/bjo.2020.160

14. Liberati E, Richards N, Parker J, Willars J, Scott D, Boydell N, Pinfold V, Martin G, Dixon-Woods M, Jones PB. Remote care for mental health: qualitative study with service users, carers and staff during the COVID19 pandemic. BMJ Open 2021;11:e049210 : doi: 10.1136/bmjopen-2021-049210

15. Dhahri AA, Iqbal MR, Pardoe H. Agile Application of Video Telemedicine During the COVID-19 Pandemic. Cureus. doi: 10.7759/cureus.11320

16. Doraiswamy S, Abraham A, Mamtani R, Cheema S. Use of Telehealth During the COVID-19 Pandemic: Scoping Review. J Med Internet Res. 2020 Dec 1;22(12):e24087. doi: 10.2196/24087.

17. Vusirikala A, Ensor D, Asokan AK, Lee AJX, Ray R, Tsekes D, Edwin J. Hello, can you hear me? Orthopaedic clinic telephone consultations in the COVID-19 era-a patient and clinician perspective. World J Orthop 2021 January 18;12(1):24–34. doi: 10.5312/wjo.v12.i1.24

18. Age UK report. How old is the UK? An Investigation into the Ageing Population of Britain. Age UK. 2018. Available from: https://www.ageukmobility.co.uk/mobility-news/article/how-old-is-the-uk (Accessed 4th December 2020).

19. Cornwall Council. Cornwall: a brief description. 2015. Available from: https://www.cornwall.gov.uk/media/20392018/cornwall-statistics-infographic-a3_proof3.pdf (Accessed 4th December 2020).

20. NHS England and NHS Improvement. Attend Anywhere. 2020 Available from: https://england.nhs.attendanywhere.com/resourcecentre/Content/Public_Topics/Discover.htm. (Accessed 3rd October 2020).

21. Remote consultations. NHS England and NHS improvement coronavirus. Specialty guides for patient management. 2020. Available from: https://www.england.nhs.uk/coronavirus/publication/specialty.guides. (Accessed 3rd October 2020).

22. NHS East Suffolk and North Essex. ‘Attend Anywhere’ Patient Feedback _ ESNEFT. 2020. Available from: https://forms.office.com/Pages/ResponsePage.aspx?id=kp4VA8ZyI0umSq9Q55Ctvy0O6tkePahBgNX4hXWu7AVUQVJNQ1g1REM4UFQwNDczUDUwTU9NTUc1My4u (Accessed 3rd December 2020).

23. Braun V, Clarke V. Using thematic analysis in psychology. Qualitative Research in Psychology. 2006;3(2):77–101. doi: 10.1191/1478088706qp063oa

24. Ogrinc G, Davies L, Goodman D, Batalden P, Davidoff F, Stevens D. SQUIRE 2.0 (Standards for Quality Improvement Reporting Excellence): revised publication guidelines from a detailed consensus process. BMJ Qual Saf. 2016 Dec;25(12):986–992. doi: 10.1136/bmjqs-2015-004411

25. Statton S, Jones R, Thomas M, North T, Endacott R, Frost A, et al. Professional learning needs in using video calls identified through workshops. BMC Medical Education. 2016;16(1):140. doi: 10.1186/s12909-016-0657-6

26. Mcgaughey R, Zeltmann S, Mcmurtrey M. Motivations and obstacles to smartphone use by the elderly: developing a research framework. International Journal of Electronic Finance. 2013;7:177–195. doi: 10.1504/IJEF.2013.058601.

27. Vaportzis E, Clausen MG, Gow AJ. Older Adults Perceptions of Technology and Barriers to Interacting with Tablet Computers: A Focus Group Study. Front Psychol. 2017;8:1687. doi:10.3389/fpsyg.2017.01687.

28. Biswas M, Romeo M, Cangelosi A, Jones R. Are older people any different from younger people in the way they want to interact with robots? Scenario based survey. Journal on Multimodal User Interfaces 2019;14(2). doi: 10.1007/s12193-019-00306-x

29. Antoni E, Jaboli M, Samarasinghe M, et al. PWE-132 Is The NHS Ready for Video-Conference Consultations in Outpatient Clinics? BMJ Gut 2016;65:A203–A204. doi: 10.1136/gutjnl-2016-312388.377

30. Age UK. Painful Journeys: Why getting to hospital appointments is a major issue for older people. 2017. Available from: In-depth policy report. https://www.ageuk.org.uk/globalassets/age-uk/documents/reports-and-publications/reports-and-briefings/active-communities/rb_dec17_painful_journeys_indepth_report.pdf (Accessed 10th December 2020).

31. Sibley M, Earwicker R, Huber J. Making best use of patient experiences. Journal of Clinical Nursing 2018;27:23–24. doi: 10.1111/jocn.14504

32. Edwards C, Staniszewska S. Accessing the user’s perspective. Health & Social Care in the Community 2000;8(6):417–424. doi: 10.1046/j.1365-2524.2000.00267.x

33. Williams B, Coyle J, Healy D. The meaning of patient satisfaction: An explanation of high reported levels. Social Science & Medicine 1998;47(9);1351-1359. doi: 10.1016/s0277-9536(98)00213-5

34. Sheard L, Peacock R, March C, Lawton R. What’s the problem with patient experience feedback? A macro and micro understanding, based on findings from a three-site UK qualitative study. Health Expectations 2019;22:46–53. doi: 10.1111/hex.12829

35. Burt J, Campbell J, Abel G, Aboulghate A, Ahmed F, Asprey A, et al. Improving patient experience in primary care: a multimethod programme of research on the measurement and improvement of patient experience. Programme Grants Appl Res. 2017;5(9). doi: 10.3310/pgfar05090

